# Abnormal spirometric patterns and respiratory symptoms in HIV patients with no recent pulmonary infection in a periurban hospital in Ghana

**DOI:** 10.1101/2022.08.02.22278341

**Authors:** Kwame Yeboah, Latif Musa, Kweku Bedu-Addo

## Abstract

**Background:** Human immunodeficiency virus (HIV) infection is associated with chronic pulmonary diseases, even in those with viral suppression by highly active antiretroviral treatment (HAART). Spirometry is an accurate method of diagnosing pulmonary dysfunction in people living with HIV (PLWH).

**Aim:** To compare the prevalence of spirometric abnormalities among HAART-treated HIV patients and HAART naïve HIV patients with non-HIV controls with no recent history of pulmonary infection in a peri-urban hospital in Ghana.

**Methods:** In a case-control design, we recruited 158 HAART-treated HIV patients, 150 HAART-naïve HIV patients and 156 non-HIV controls for the study. Clinical, sociodemographic data and respiratory symptoms were collected using a structured questionnaire. Spirometry was performed in all participants and abnormalities were categorised as obstructive (OSP) or restrictive (RSP) spirometric patterns based on the GLI definition.

**Results:** The prevalence of OSP was similar among the HAART treated, HAART naïve HIV patients and non-HIV controls (10.1% vs 9.3% vs 9% respectively, p=0.994), whereas that of RSP was higher in HAART-treated HIV patients compared to HAART-naïve HIV patients and non-HIV controls (51.9% vs 32.1% vs 32% respectively, p=0.013). Respiratory symptoms were common among HAART-treated and HAART-naïve HIV patients compared to non-HIV controls (48.1% vs 40% vs 19.2% respectively, p<0.001). The major determinants of OSP were female gender, exposure to medium-to-high levels of biomass, presence of a respiratory symptom, unemployment and underweight, and that of RSP were age, female gender, being unmarried, medium-to-high biomass exposure and being self-employed or unemployed.

**Conclusion:** In HIV patients without any recent pulmonary infection in a peri-urban area of Ghana, there was no difference in the prevalence of OSP among HAART-treated and HAART naïve HIV patients compared to the non-HIV control. However, the prevalence of RSP was higher in HAART-treated HIV patients compared to the other groups.

## Introduction

Sub-Saharan Africa (SSA) has the largest population of people living with human immunodeficiency virus (HIV) and this is associated with healthcare, socio-economic and developmental challenges [1]. It was reported in 2017 that 71% of the global total number of people living with HIV (PLWH) resides in SSA, with 75% of deaths and 65% of new HIV infections occurring in the region [2]. In Ghana, the prevalence of HIV infection is 1.7% and is concentrated mostly in urban areas [3]. With the introduction of effective highly active antiretroviral therapy (HAART), PLWH now live longer due to decreased morbidity and mortality from opportunistic infections [4]. This has increased the susceptibility of PLWH to chronic diseases at greater rates than those observed in HIV-uninfected persons, making HIV infection a manageable chronic disease [5]. Several studies have reported a high burden of chronic lung diseases in PLWH, even after adjustments for risk factors such as smoking, opportunistic infections and injection drug use [4, 6-9].

Spirometry is a simple, effective and reproducible method for diagnosing chronic lung diseases, and the results can be categorized into two major patterns of abnormalities. An obstructive spirometric pattern (OSP) is characterized by a disproportional reduction in the forced expiratory volume in 1 s (FEV1) relative to the forced vital capacity (FVC), leading to a reduction in the FEV1/FVC ratio to less than the lower limit of normal (LLN) of a similar population [10, 11]. Restrictive spirometric pattern (RSP) is defined by a normal FEV1/FVC (>LLN), but an FVC is less than 80% of the predicted value. Both obstructive and restrictive spirometry patterns are associated with increased mortality in the general population [11, 12]. Despite the high prevalence of PLWH in SSA, few studies have reported the prevalence and risk factors for these abnormal spirometric patterns with recent reports from the FRESH AIR study in Uganda [5, 6, 13-17]. Most of these studies attribute abnormal spirometry in HIV to opportunistic pulmonary infection. We compared the prevalence of spirometric abnormalities among HIV patients on HAART treatment and HAART-naïve HIV patients with non-HIV controls with no recent history of pulmonary infection in a peri-urban hospital in Ghana. We hypothesize that HIV infection would increase the prevalence of spirometric abnormalities compared to those with no HIV infection.

## Methods

### Study participants and design

This study was a case-control design with HIV patients as cases and the controls were non-HIV individuals who visited the facility for voluntary testing of their HIV status. HIV patients were categorized as those on HAART management and HAART-naïve HIV patients who have not been on HAART treatment for more than 4 weeks. The study was conducted at Atua Government Hospital, a 150-bed primary healthcare facility, located in Agormanya, a periurban town in the Eastern region of Ghana. The Agormanya area has a higher prevalence of HIV infection (11.6%) compared to the national prevalence which stands at 1.7% in 2020 [3]. The hospital has about 1,500 HIV patients on its register. Participants with conditions contraindicated for spirometry, diagnosis of lung infection or disease from their clinical case notes or answering positively to the question: “Have you been told by your health worker that you have lung disease in the past years?” were excluded from the study. Ethical approval was obtained from the College of Health Sciences’ Ethical & Protocol Review Committee and all participants provided voluntary informed consent before joining the study.

### Data collection

A structured questionnaire was used to obtain data on sociodemographic factors like age, gender, lifestyle factors (smoking, alcohol intake), medical history (hypertension, diabetes, cardiovascular disease), current medication (pulmonary medication, antihypertensive agents, anti-diabetic drugs), occupation, education, marital status and biomass exposure. Medium-high biomass exposure was defined as those who reported using firewood, sawdust, cow-dung or corn cubs as a fuel source for work or cooking at home on most days for more than 6 months. Respiratory symptoms were assessed using a modified version of the American Thoracic Society respiratory questionnaire, which included the following questions: “Do you usually have coughs without cold for more than 2 months in a year?”, “Do you usually bring up phlegm from your chest when you don’t have a cold?”, “Do you have difficulty breathing when you walk fast, use a staircase or run?” and “Have you ever had an attack of wheezing or whistling in your chest that made you feel short of breath?” [18].

Body weight and height were measured using a stadiometer in light clothing with footwear removed, with body mass index (BMI) calculated as weight/height^2^ and categorized as: underweight (< 18.5 kg/m^2^), normal weight (18.5–24.9 kg/m^2^), overweight (25–29.9 kg/m^2^), and obese (≥ 30 kg/m^2^). Hypertension was defined as self-reported use of antihypertensive treatment and/or systolic blood pressure ≥ 140 mm Hg and/or diastolic blood pressure ≥ 90 mm Hg. We collected blood samples and measured the levels of CD4+ lymphocytes at the time of the study.

Spirometry was performed by a qualified technician using a Vitalograph Pneumotrac desktop spirometer model 6800 following the ATS/ERS 2019 standard guidelines [19]. Using the Global Lung Initiative (GLI) 2012 reference equations for African Americans, normal lung function pattern was defined by FEV1, FVC, and FEV1/FVC all > LLN. Obstructive lung function was determined by the ratio of Forced expiratory volume in the first second (FEV1) to Forced Vital Capacity (FVC) that was below the LLN with Forced Vital Capacity above LLN (FEV1/FVC <LLN with FVC > LLN), while restrictive lung function abnormalities were suggested by values less than the LLN for the FVC (FVC < LLN with FEV1/FVC > LLN) [20].

### Sample size

The sample size required for the studies was calculated based on the prevalence of COPD reported in the rural Ugandan population to be 16.2% [21]. With the assumption that the prevalence ratio in the HIV population would be 2.2, we will need at least 147 participants in each group to achieve a power of 80% within a 95% significance level.

### Data analysis

Data were analysed using IBM SPSS version 28 software. Continuous descriptive data were presented as means and standard deviations for normally distributed data and comparisons between groups were made using one-way ANOVA, while non-normally distributed data were presented as medians and interquartile ranges and comparisons were made using the Kruskal-Wallis’ test. Categorical descriptive data were presented as frequencies and percentages and analysed using the chi-square test. Univariate and multivariable logistic regression models were used to analyse the change in odds of abnormal spirometry and respiratory symptoms with socio-demographic and clinical factors. p< 0.05 was considered statistically significant.

## Results

### General characteristics of study participants

The age range of participants was 20 – 70 years with a mean age of 38.4±13.7 years. There was no difference in mean age among various categories of participants. There was a high proportion of females in this study participants and participants who currently or formerly smoked were mostly HIV patients. HIV patients were likely to be unemployed or self-employed and had no formal education or attended school up to junior high level. HIV patients had greater biomass exposure from work or home and were more likely to be underweight. Measurements from spirometry indicate that HAART-treated HIV patients had reduced FEV_1_, FVC and FEF_25 – 75_ l/min compared to HAART-naïve HIV patients and non-HIV participants. HAART-treated HIV patients had a high prevalence of RSP compared to HAART-naïve HIV patients and non-HIV controls, but there was no difference in the prevalence of OSP or undefined pulmonary patterns among various groups of participants (Table 1).

**Table 1.**
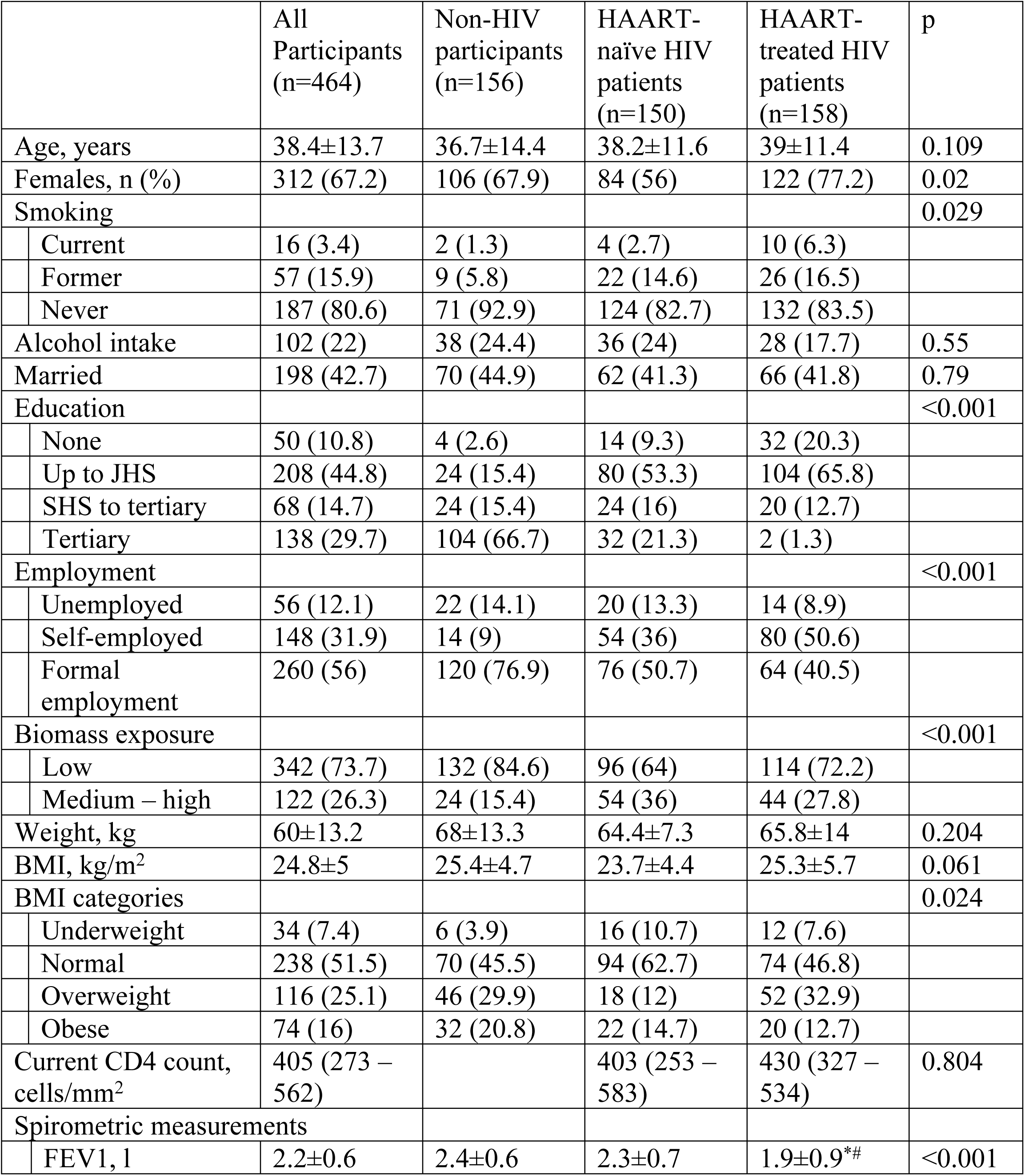
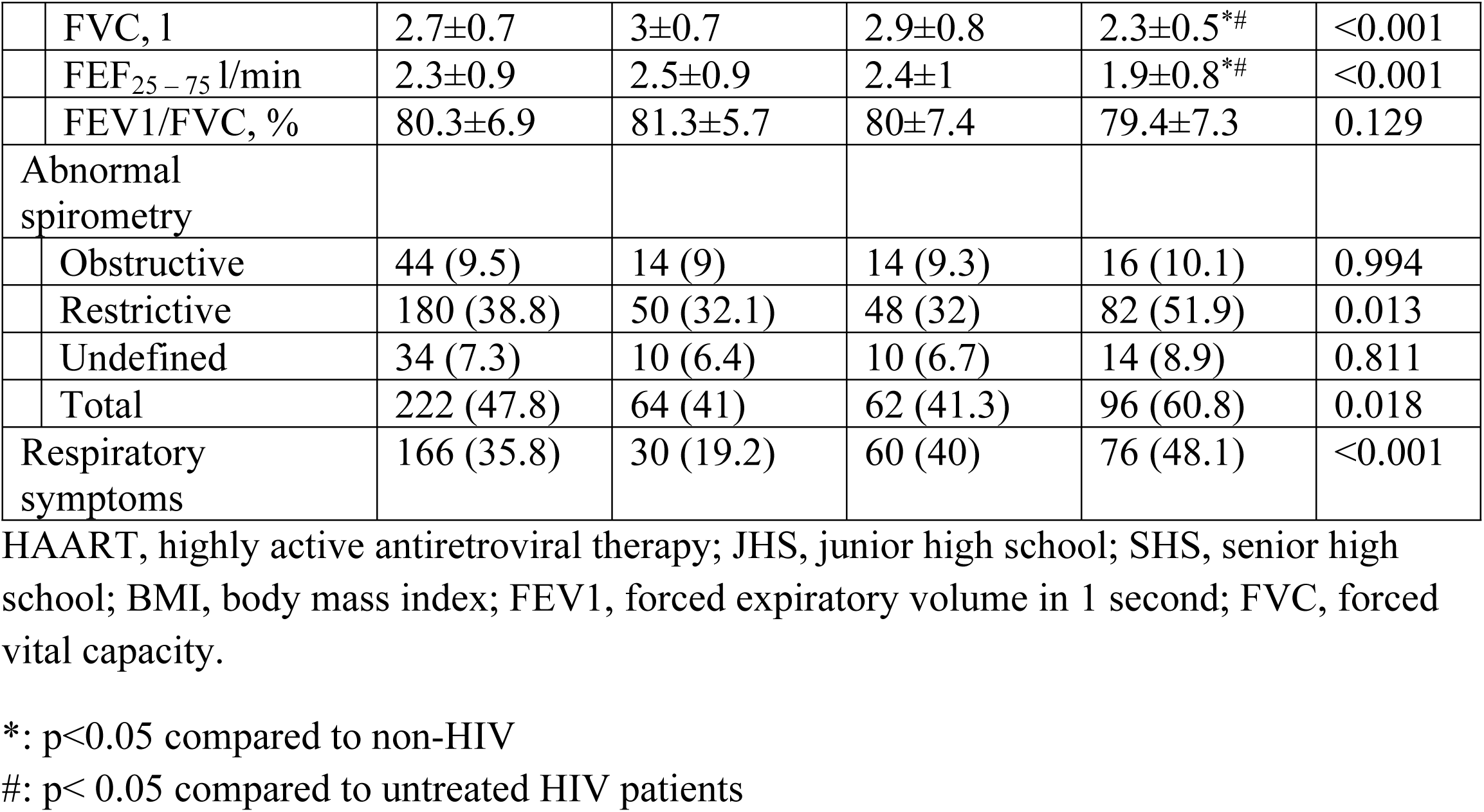
General characteristics of study participants

|  | All Participants (n=464) | Non-HIV participants (n=156) | HAART-naïve HIV patients (n=150) | HAART-treated HIV patients (n=158) | p |
| --- | --- | --- | --- | --- | --- |
| Age, years | 38.4±13.7 | 36.7±14.4 | 38.2±11.6 | 39±11.4 | 0.109 |
| Females, n (%) | 312 (67.2) | 106 (67.9) | 84 (56) | 122 (77.2) | 0.02 |
| Smoking |  |  |  |  | 0.029 |
| Current | 16 (3.4) | 2 (1.3) | 4 (2.7) | 10 (6.3) |  |
| Former | 57 (15.9) | 9 (5.8) | 22 (14.6) | 26 (16.5) |  |
| Never | 187 (80.6) | 71 (92.9) | 124 (82.7) | 132 (83.5) |  |
| Alcohol intake | 102 (22) | 38 (24.4) | 36 (24) | 28 (17.7) | 0.55 |
| Married | 198 (42.7) | 70 (44.9) | 62 (41.3) | 66 (41.8) | 0.79 |
| Education |  |  |  |  | <0.001 |
| None | 50 (10.8) | 4 (2.6) | 14 (9.3) | 32 (20.3) |  |
| Up to JHS | 208 (44.8) | 24 (15.4) | 80 (53.3) | 104 (65.8) |  |
| SHS to tertiary | 68 (14.7) | 24 (15.4) | 24 (16) | 20 (12.7) |  |
| Tertiary | 138 (29.7) | 104 (66.7) | 32 (21.3) | 2 (1.3) |  |
| Employment |  |  |  |  | <0.001 |
| Unemployed | 56 (12.1) | 22 (14.1) | 20 (13.3) | 14 (8.9) |  |
| Self-employed | 148 (31.9) | 14 (9) | 54 (36) | 80 (50.6) |  |
| Formal employment | 260 (56) | 120 (76.9) | 76 (50.7) | 64 (40.5) |  |
| Biomass exposure |  |  |  |  | <0.001 |
| Low | 342 (73.7) | 132 (84.6) | 96 (64) | 114 (72.2) |  |
| Medium – high | 122 (26.3) | 24 (15.4) | 54 (36) | 44 (27.8) |  |
| Weight, kg | 60±13.2 | 68±13.3 | 64.4±7.3 | 65.8±14 | 0.204 |
| BMI, kg/m<sup>2</sup> | 24.8±5 | 25.4±4.7 | 23.7±4.4 | 25.3±5.7 | 0.061 |
| BMI categories |  |  |  |  | 0.024 |
| Underweight | 34 (7.4) | 6 (3.9) | 16 (10.7) | 12 (7.6) |  |
| Normal | 238 (51.5) | 70 (45.5) | 94 (62.7) | 74 (46.8) |  |
| Overweight | 116 (25.1) | 46 (29.9) | 18 (12) | 52 (32.9) |  |
| Obese | 74 (16) | 32 (20.8) | 22 (14.7) | 20 (12.7) |  |
| Current CD4 count, cells/mm<sup>2</sup> | 405 (273 – 562) |  | 403 (253 – 583) | 430 (327 – 534) | 0.804 |
| Spirometric measurements |  |  |  |  |  |
| FEV<sub>1</sub>, l | 2.2±0.6 | 2.4±0.6 | 2.3±0.7 | 1.9±0.9<sup>*#</sup> | <0.001 |

|  |  |  |  |  |  |  |
| --- | --- | --- | --- | --- | --- | --- |
|  | FVC, l | 2.7±0.7 | 3±0.7 | 2.9±0.8 | 2.3±0.5*# | <0.001 |
|  | FEF<sub>25-75</sub> l/min | 2.3±0.9 | 2.5±0.9 | 2.4±1 | 1.9±0.8*# | <0.001 |
|  | FEV1/FVC, % | 80.3±6.9 | 81.3±5.7 | 80±7.4 | 79.4±7.3 | 0.129 |
| Abnormal spirometry |  |  |  |  |  |  |
|  | Obstructive | 44 (9.5) | 14 (9) | 14 (9.3) | 16 (10.1) | 0.994 |
|  | Restrictive | 180 (38.8) | 50 (32.1) | 48 (32) | 82 (51.9) | 0.013 |
|  | Undefined | 34 (7.3) | 10 (6.4) | 10 (6.7) | 14 (8.9) | 0.811 |
|  | Total | 222 (47.8) | 64 (41) | 62 (41.3) | 96 (60.8) | 0.018 |
| Respiratory symptoms |  | 166 (35.8) | 30 (19.2) | 60 (40) | 76 (48.1) | <0.001 |

HAART, highly active antiretroviral therapy; JHS, junior high school; SHS, senior high school; BMI, body mass index; FEV1, forced expiratory volume in 1 second; FVC, forced vital capacity.
\*: $p < 0.05$ compared to non-HIV
#: $p < 0.05$ compared to untreated HIV patients

### Respiratory symptoms

Among the entire study participants, 35.8% reported the presence of at least one respiratory symptom, with a high proportion in HAART-treated and HAART-naïve HIV patients compared to non-HIV participants (48.1% vs 40% vs 19.2 respectively, p<0.001). Phlegm production and regular cough were commonly reported in 26.4% and 19% of the entire study participants respectively, with high proportion in HAART-treated and HAART-naïve HIV patients compared to non-HIV participants (phlegm: 35.4% vs 26.7% vs 6.4%, p<0.001; regular coughs: 30.4% vs 20% vs 11.5%, p<0.001 respectively). Similar trends were observed in participants who reported having dyspnoea on exertion and frequent wheeze symptoms (Fig. 1). Increasing age, being unemployed, exposure to a high-medium level of biomass, being HIV patients with or without HAART treatment and having basic or no education was associated with increased odds of having respiratory symptoms in unadjusted regression models. In the adjusted regression models, exposure to medium-high level of biomass, being HAART treated HIV or HAART-naïve HIV patient were associated with increased odds of having respiratory symptoms (Table 2).

**Table 2.** Factors associated with respiratory symptoms from logistic regression analyses

|  |  | Unadjusted model |  | Adjusted model |  |
| --- | --- | --- | --- | --- | --- |
|  |  | OR (95% CI) | p | OR (95% CI) | p |
| Age |  | 1.02 (1.01 – 1.03) | 0.007 | 1 (0.98 – 1.02) | 0.734 |
| Females |  | 1.32 (0.87 – 1.99) | 0.189 | 1.14 (0.69 – 1.88) | 0.623 |
| Unmarried |  | 1.34 (0.91 – 1.97) | 0.14 | 1.18 (0.76 – 1.86) | 0.462 |
| Alcohol intake |  | 0.97 (0.62 – 1.54) | 0.909 | 1 (0.59 – 1.68) | 0.986 |
| Current smoking |  | 1.84 (0.68 – 4.98) | 0.233 | 1.52 (0.5 – 4.64) | 0.463 |
| Medium/high biomass exposure |  | 2.72 (1.12 – 4.57) | 0.008 | 1.87 (1.05 – 3.49) | 0.017 |
| HIV status: reference= non-HIV |  |  |  |  |  |
|  | Treatment naïve | 2.8 (1.67 – 4.69) | <0.001 | 2.03 (1.1 – 3.74) | 0.023 |
|  | HAART treatment | 3.89 (2.35 – 6.46) | <0.001 | 2.61 (1.32 – 5.17) | 0.006 |
| Employment |  |  |  |  |  |
|  | Self-employed | 1.4 (0.92 – 2.13) | 0.118 | 0.83 (0.5 – 1.36) | 0.451 |
|  | Unemployed | 1.88 (1.05 – 3.38) | 0.035 | 1.78 (0.91 – 3.48) | 0.091 |
| Education: reference= tertiary |  |  |  |  |  |
|  | None | 3.32 (1.67 – 6.6) | <0.001 | 2.22 (0.89 – 5.51) | 0.086 |
|  | Basic/JHS | 3.21 (1.97 – 5.22) | <0.001 | 1.79 (0.89 – 3.62) | 0.102 |
|  | SHS/Tech | 0.93 (0.46 – 1.91) | 0.85 | 0.62 (0.27 – 1.43) | 0.263 |
| BMI (unit change) |  | 1 (0.95 – 1.03) | 0.587 |  |  |
| BMI categories |  |  |  |  |  |
|  | Underweight | 1.85 (0.9 – 3.81) | 0.095 | 1.58 (0.73 – 3.42) | 0.25 |
|  | Overweight | 0.68 (0.42 – 1.1) | 0.116 | 0.6 (0.35 – 1.03) | 0.062 |
|  | Obese | 0.79 (0.45 – 1.37) | 0.789 | 0.82 (0.44 – 1.51) | 0.517 |

HAART, highly active antiretroviral therapy; JHS, junior high school; SHS, senior high school; BMI, body mass index.

**Fig 1.**
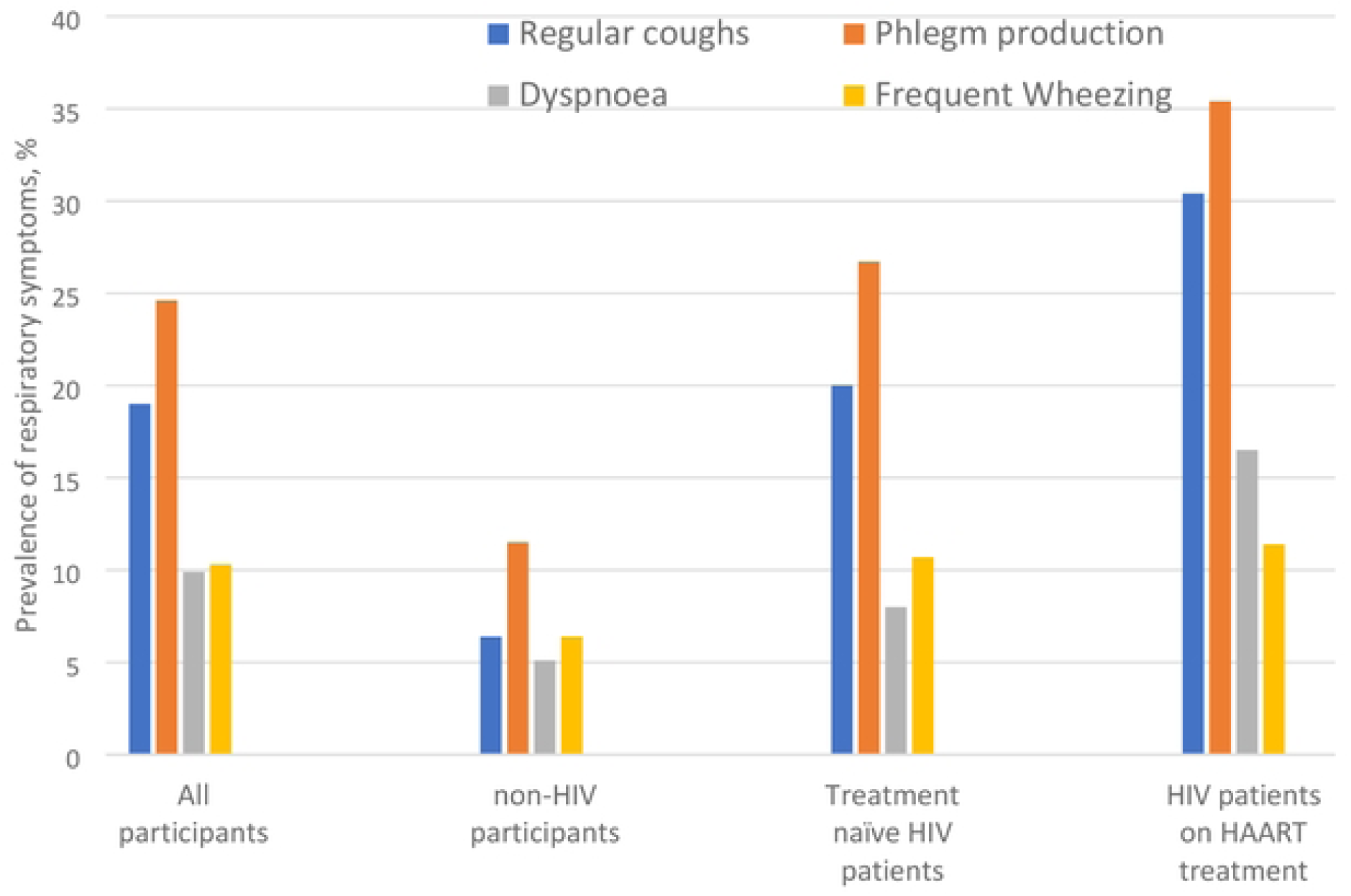
Prevalence of respiratory symptoms in study participants by the HIV status

### Determinants of abnormal spirometric patterns

In logistic regression analyses, being female, unmarried, unemployed, having no formal education, underweight and having a respiratory symptom were associated with increased odds of having OSP in the unadjusted models. In the adjusted models, female gender, medium-to-high biomass exposure, presence of respiratory symptoms, unemployed and underweight were associated with increased odds of having OSP (Table 3). The determinant of OSP among various categories of participants based on HIV status is shown in the supplementary attachment (Table S1 – S3).

**Table 3.**
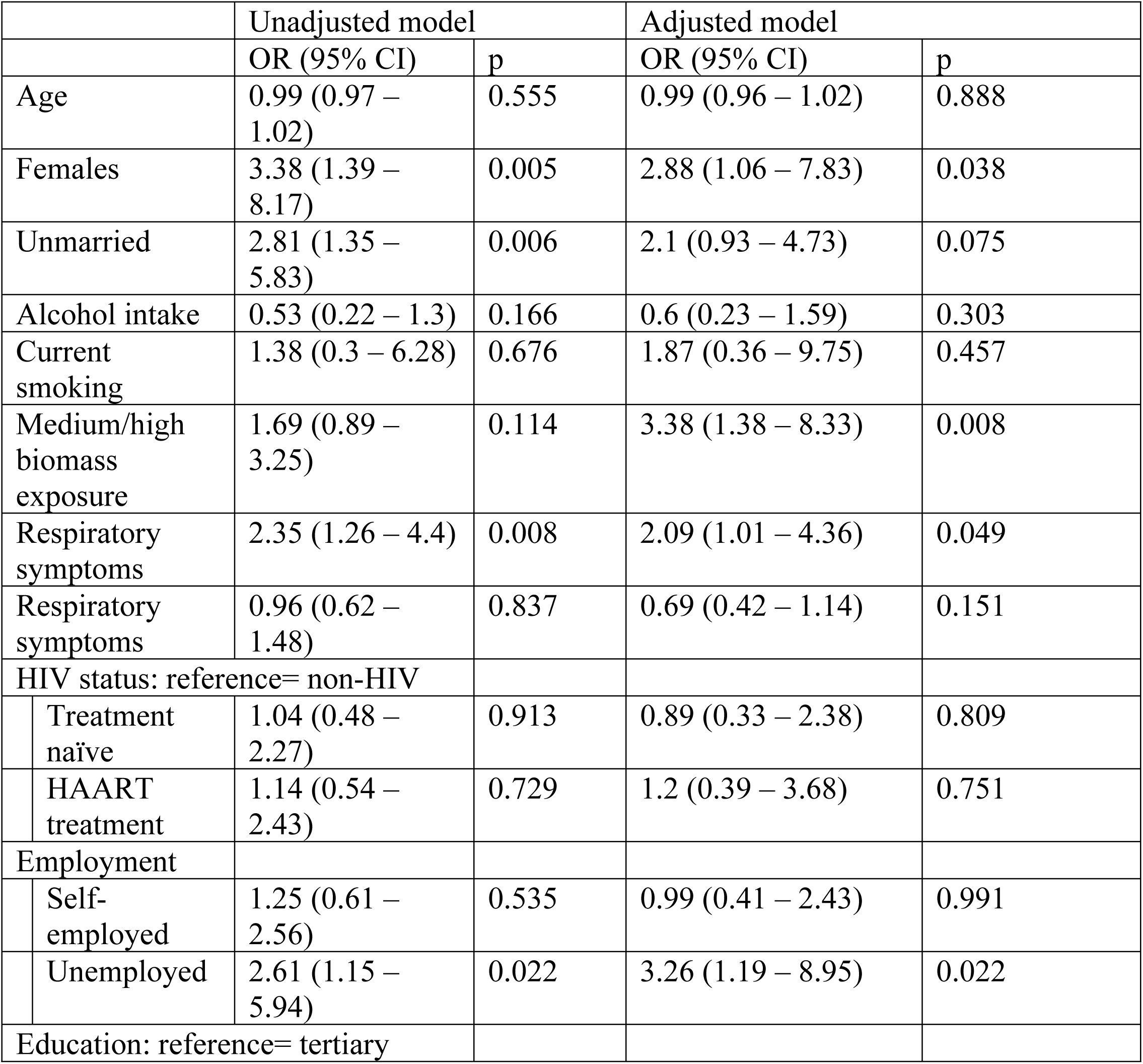
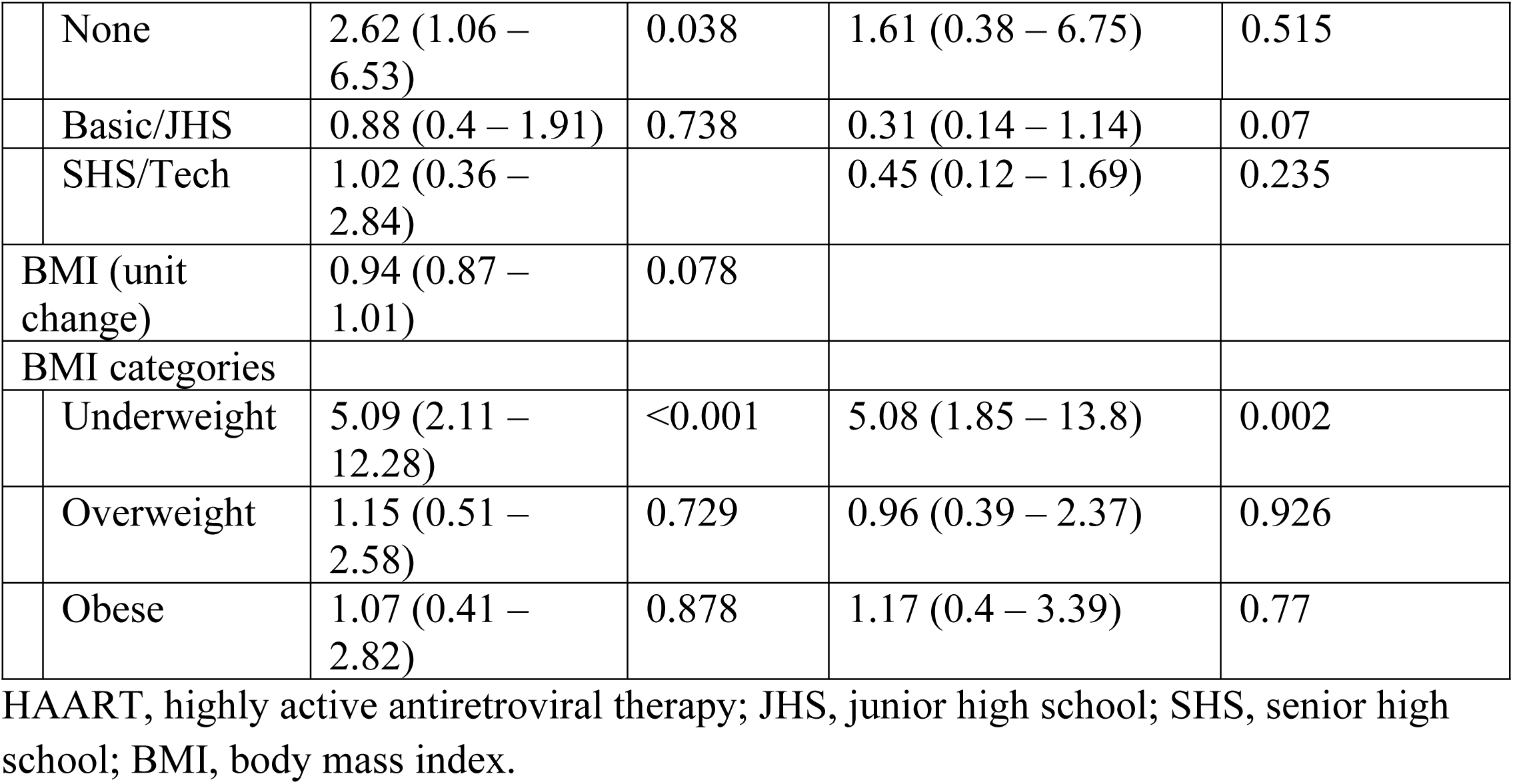
Factors associated with obstructive spirometric pattern from logistic regression analyses

Concerning RSP, female gender, medium-to-high biomass exposure and being HAART-treated HIV patient were associated with increased odds in unadjusted logistic regression models. In multivariable logistic regression models, age, female gender, being unmarried, medium-to-high biomass exposure and being self-employed or unemployed were associated with increased odds of restrictive lung pattern (Table 4). The determinants of RSP among various patient groups are shown in the supplementary attachment (Tables S4 – S6).

**Table 4.**
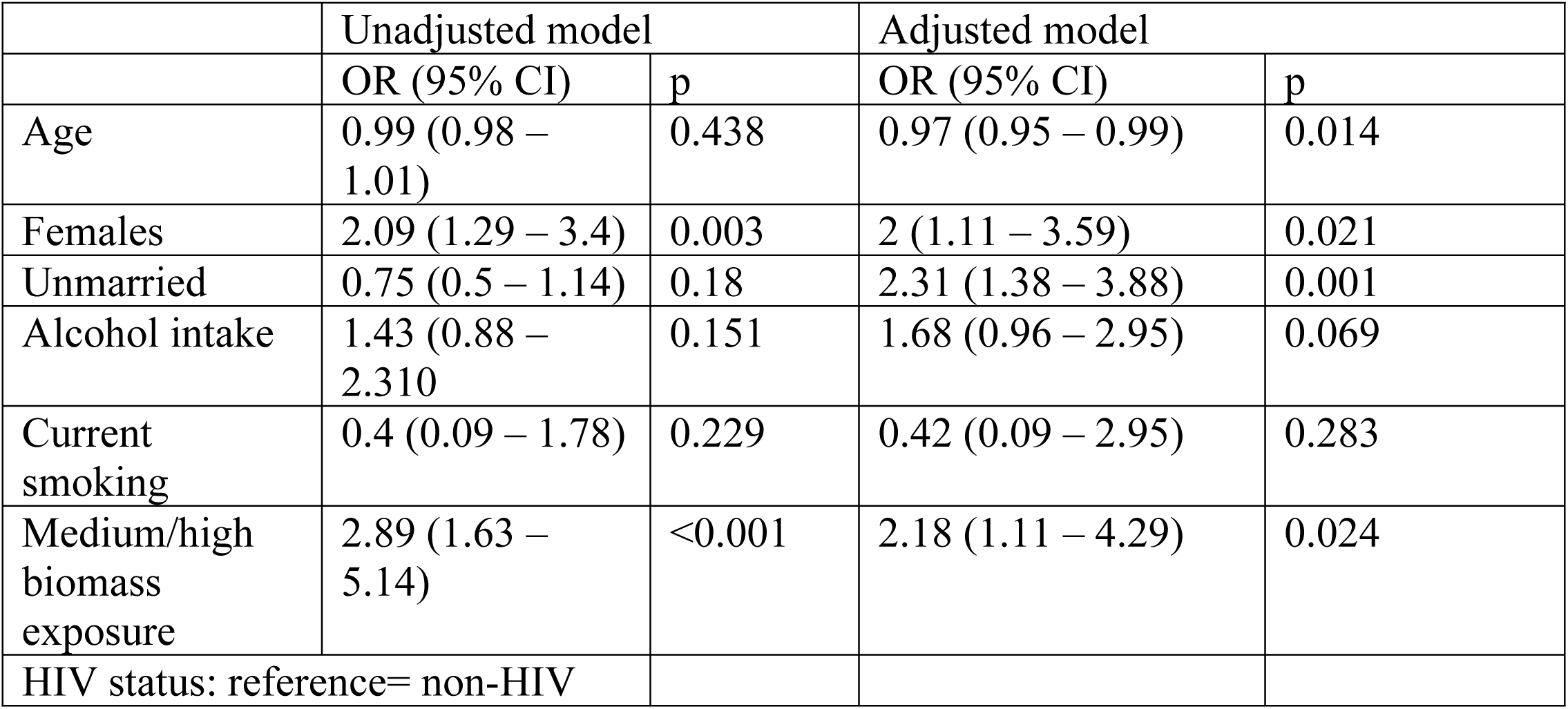
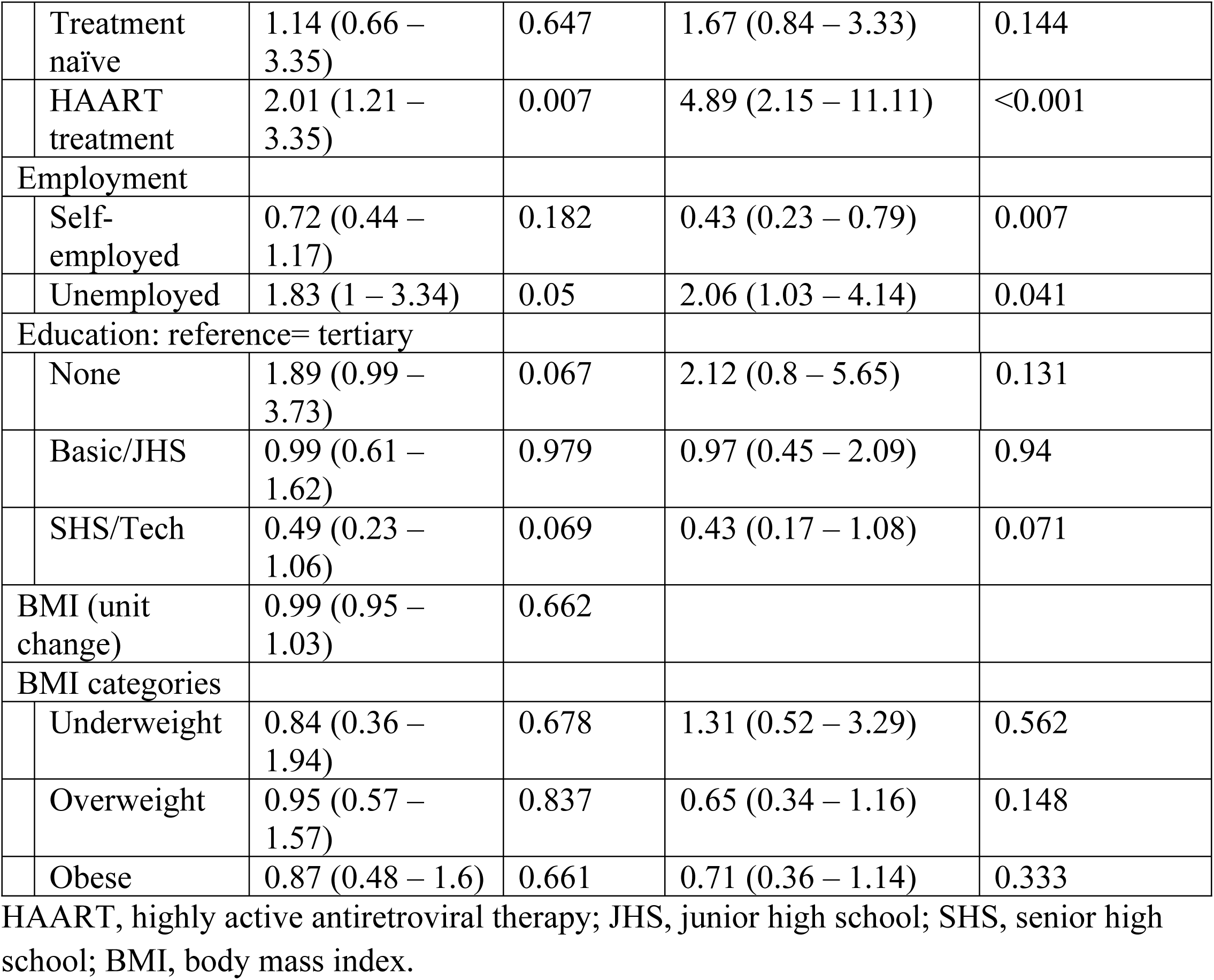
Factors associated with restrictive spirometric pattern from logistic regression analyses

|  | Unadjusted model |  | Adjusted model |  |
| --- | --- | --- | --- | --- |
|  | OR (95% CI) | p | OR (95% CI) | p |
| Age | 0.99 (0.98 – 1.01) | 0.438 | 0.97 (0.95 – 0.99) | 0.014 |
| Females | 2.09 (1.29 – 3.4) | 0.003 | 2 (1.11 – 3.59) | 0.021 |
| Unmarried | 0.75 (0.5 – 1.14) | 0.18 | 2.31 (1.38 – 3.88) | 0.001 |
| Alcohol intake | 1.43 (0.88 – 2.310) | 0.151 | 1.68 (0.96 – 2.95) | 0.069 |
| Current smoking | 0.4 (0.09 – 1.78) | 0.229 | 0.42 (0.09 – 2.95) | 0.283 |
| Medium/high biomass exposure | 2.89 (1.63 – 5.14) | <0.001 | 2.18 (1.11 – 4.29) | 0.024 |
| HIV status: reference= non-HIV |  |  |  |  |

|  |  |  |  |  |  |
| --- | --- | --- | --- | --- | --- |
|  | Treatment naïve | 1.14 (0.66 – 3.35) | 0.647 | 1.67 (0.84 – 3.33) | 0.144 |
|  | HAART treatment | 2.01 (1.21 – 3.35) | 0.007 | 4.89 (2.15 – 11.11) | <0.001 |
| Employment |  |  |  |  |  |
|  | Self-employed | 0.72 (0.44 – 1.17) | 0.182 | 0.43 (0.23 – 0.79) | 0.007 |
|  | Unemployed | 1.83 (1 – 3.34) | 0.05 | 2.06 (1.03 – 4.14) | 0.041 |
| Education: reference= tertiary |  |  |  |  |  |
|  | None | 1.89 (0.99 – 3.73) | 0.067 | 2.12 (0.8 – 5.65) | 0.131 |
|  | Basic/JHS | 0.99 (0.61 – 1.62) | 0.979 | 0.97 (0.45 – 2.09) | 0.94 |
|  | SHS/Tech | 0.49 (0.23 – 1.06) | 0.069 | 0.43 (0.17 – 1.08) | 0.071 |
| BMI (unit change) |  | 0.99 (0.95 – 1.03) | 0.662 |  |  |
| BMI categories |  |  |  |  |  |
|  | Underweight | 0.84 (0.36 – 1.94) | 0.678 | 1.31 (0.52 – 3.29) | 0.562 |
|  | Overweight | 0.95 (0.57 – 1.57) | 0.837 | 0.65 (0.34 – 1.16) | 0.148 |
|  | Obese | 0.87 (0.48 – 1.6) | 0.661 | 0.71 (0.36 – 1.14) | 0.333 |

HAART, highly active antiretroviral therapy; JHS, junior high school; SHS, senior high school; BMI, body mass index.

### Association between respiratory symptoms and abnormal spirometric patterns

In the entire study participants with OSP, 14 (31.8%) reported having regular cough, 16 (36.4%) reported of regular phlegm production, and 12 (16.7%) reported frequent wheezing or dyspnoea on exertion. Among the participants with RSP, those reporting regular cough or frequent wheeze or dyspnoea on exertion were 24 (16.7) while 34 (23.6%) reported regular phlegm production. In logistic regression analyses, regular cough, frequent wheeze and dyspnoea on exertion were associated with increased odds of having OSP in crude and adjusted models (Fig. 2). Participants with RSP had increased odds of frequent wheeze and dyspnoea on exertion in unadjusted logistic regression analysis, but there was no association between respiratory symptoms and RSP in multivariable analyses (Fig. 3).

**Fig 2.**
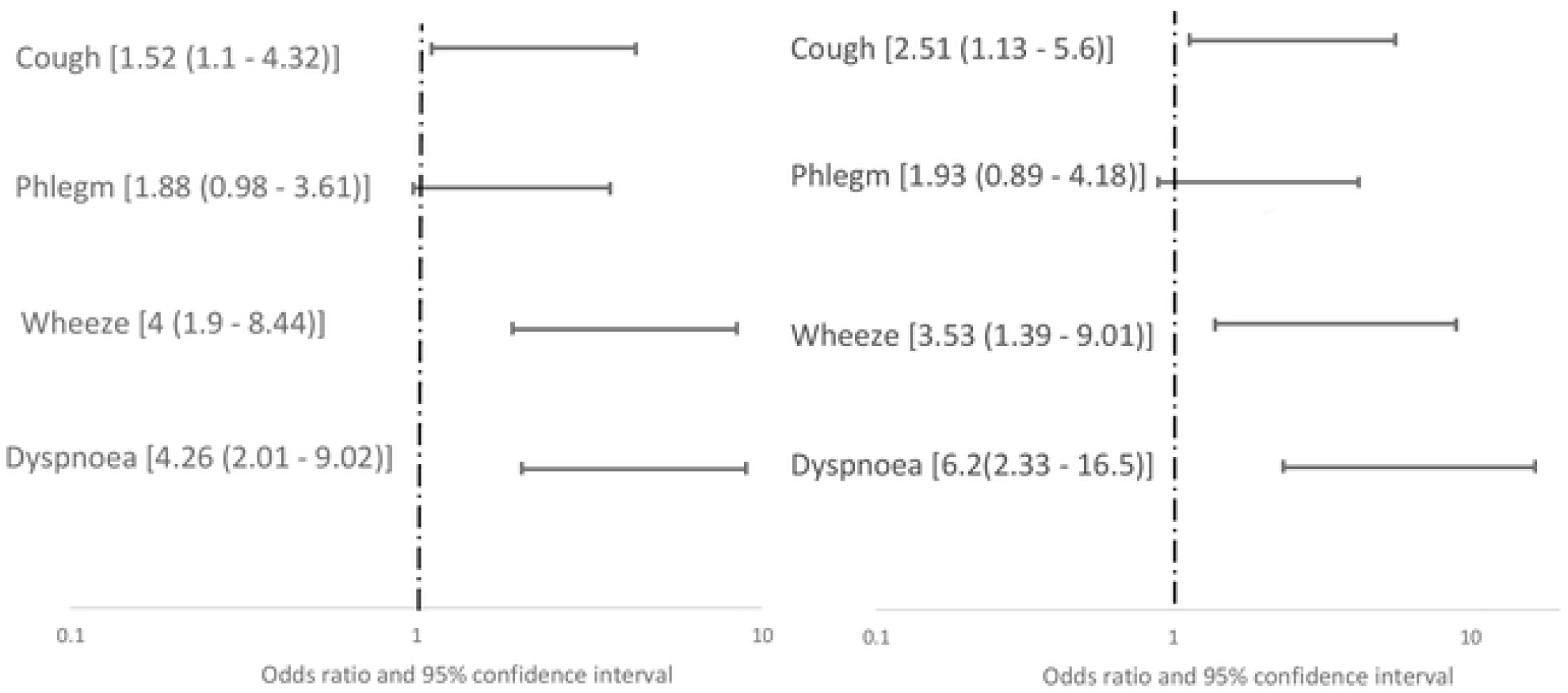
Association between respiratory symptoms and obstructive pulmonary pattern from unadjusted (left) and adjusted (right) logistic regression models.

**Fig 3.**
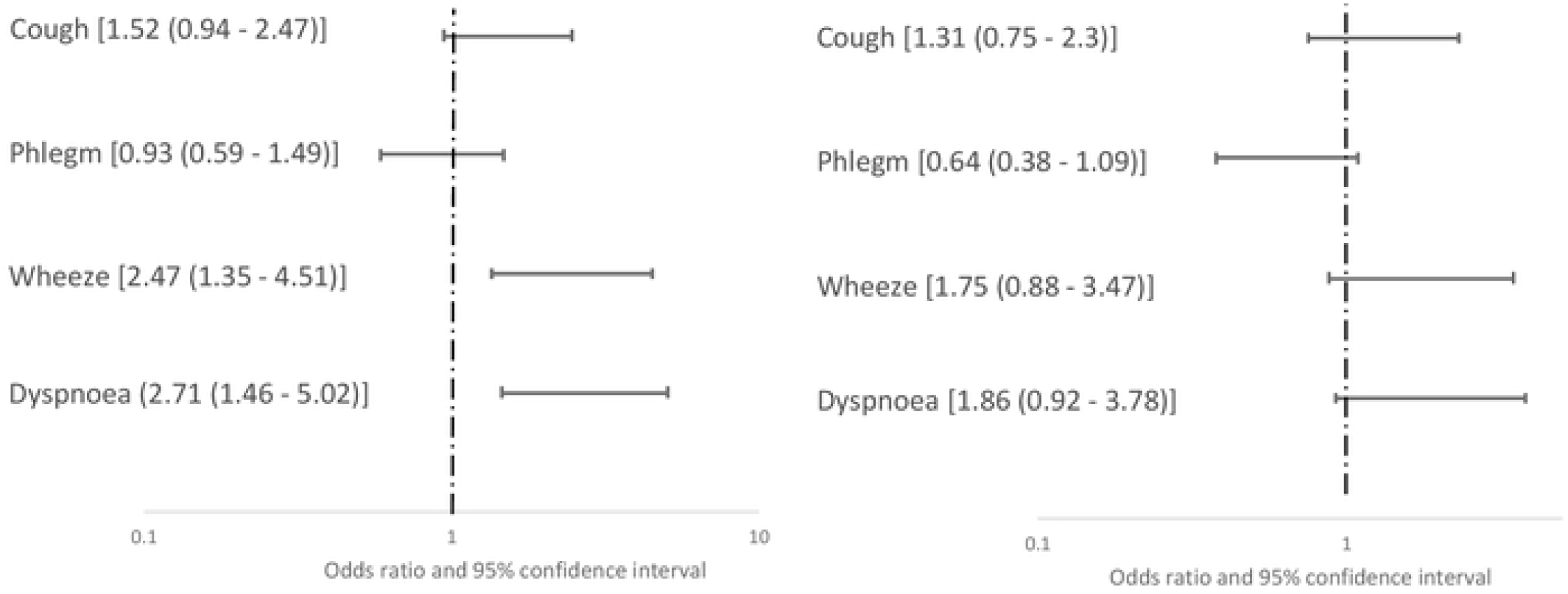
Association between respiratory symptoms and restrictive pulmonary pattern from unadjusted (left) and adjusted (right) logistic regression models.

## Discussion

### Major findings

This is the first study to report the prevalence of abnormal spirometry in HIV patients in Ghana. The major findings of the study were 1) The prevalence of OSP was similar among the HIV patient groups and controls, whereas the prevalence of RSP is higher in HIV patients on HAART treatment compared to treatment naïve HIV patients and non-HIV controls, 2) respiratory symptoms were common among HIV patients compared to non-HIV controls, and 3) the major determinants of OSP were female gender, exposure to medium-to-high levels of biomass, presence of a respiratory symptom, unemployment and underweight and that of RSP were age, female gender, being unmarried, medium-to-high biomass exposure and being self-employed or unemployed.

### Impaired spirometric measurement

We did not find any difference in the prevalence of OSP among the HIV patient groups and non-HIV controls. Few studies have compared the prevalence of OSP in HAART-treated HIV patients to that of HAART-naïve HIV patients and non-HIV controls. In the South African population, van Riel *et al* reported a higher prevalence of OSP in HAART-treated HIV patients (34.2%) compared to HAART-naïve HIV patients (11.2%). However, they used a cut-off of 20%-LLN instead of the globally accepted 5%-LLN, that was used to characterize OSP in our study. This may explain the disparity in the prevalence of OSP in our study with that of van Riel *et al* [17]. In agreement with our study, Pefura-Yone *et al* reported that there was no difference in the prevalence of OSP between HIV patients and non-HIV controls using the LLN cut-off. However, the prevalence of OSP in their HIV patients and non-HIV controls were lower than what is reported in our study [16]. Other cross-sectional studies in HIV patients in sub-Saharan Africa have reported OSP prevalence between 3.1 – 15.4% [9, 13-17], and a metanalysis reported the prevalence of OSP in HIV patients from 30 observational studies involving 151, 686 participants around the globe to be between 5.6% and 10.6% [6], similar to what was found in this study.

We found that the prevalence of RSP was similar among HAART-naïve HIV patients and non-HIV controls, but more common in HAART-treated HIV patients. Most spirometry studies conducted in HIV patients and other populations in SSA do not report the prevalence and associated factors of RSP, limiting our ability to compare the prevalence of RSP found in our study to other spirometry studies in SSA. However, Drummond *et al* reported in HIV patients in the United States that, out of 37% of HIV patients who had abnormal spirometry, 10% had RSP while 27% had OSP [8], a pattern contrary to what was observed in our study population with RSP higher than OSP. The prevalence of RSP observed in our non-HIV controls (32.1%) was similar to 36% reported by Schultz *et al* in Brazilians [22], but very high compared to 7 – 14% reported in the general US population [8], 5.9% reported in the older population [23] and 4.7% reported in Peruvian adults [24]. The accuracy of the definition of RSP used in this and many other studies, FVC<80% predicted and FEV_1_/FVC>LLN, in diagnosing RSP has been reported to be poor with low sensitivity but high negative predictive value to exclude patients without restrictive lung disease [25]. On the other hand, some study reported that using FVC<80% predicted alone, without FEV_1_/FVC>LLN, improves specificity and negative prediction of diagnosing restrictive lung disease [11]. These inconsistent definitions of RSP in literature and the failure of current guidelines to address this have resulted in the underappreciation of RSP values in spirometric studies. However, the utility and importance of RSP have been highlighted in some studies as a predictor of cardiovascular morbidity and mortality. For instance, the Cardiovascular Health Study reported that RSP is a significant predictor of CVD mortality and non-CVD mortality in older Caucasians [25]. Similarly, the TESOAD study reported that, although consistent RSP is found in 50% of patients with one-time spirometry, it is associated with an increased risk of all-cause mortality, and specifically, an increased risk of dying from diabetes and heart diseases [12]. Therefore, there is a need for further investigation into the relevance of high burden of RSP found in HIV patients and non-HIV controls in our study to characterize them properly to prevent future CVD events and deaths.

### Determinants of abnormal spirometry

The female gender was associated with an increased likelihood of having abnormal spirometry in our study. This may be because of a high proportion of females in our study participants (67.2%). A systematic review has shown that females are more likely to report asthma and airway obstruction compared to men, but abnormal spirometry is common in both genders [26, 27]. Unlike many studies that reported an association between cigarette smoking and airway obstruction, we did not find any association between current smoking and abnormal spirometry, possibly due to the low proportion of smokers in all our study participants (3.4%), but higher in HIV patient groups. The prevalence of smoking in our study is similar to that of the general Ghanaian population (4.85%) as reported by the 2014 Ghana Demographic Health Survey [28]. However, in agreement with the findings of other studies, biomass exposure was associated with abnormal spirometry in our participants [13, 15]. Biomass fumes contain about 50 air pollutants, with significant exposure similar to regular cigarette smoking [29]. These pollutants can irritate the respiratory tract by causing inflammation and tissue injury such as the excessive release of matrix metalloproteinases and increased hyaluronidase activity [30] that damage the lungs. We did not find any association between the HAART regimen used, CD4 cell count and abnormal spirometry, similar to what was reported in other HIV studies [8, 13, 15].

### Respiratory symptoms

The prevalence of respiratory symptoms in our HIV population is similar to what has been reported in other sub-Saharan African populations. In urban Ugandan HIV patients, Ddungu *et al* reported the prevalence of any respiratory symptom to be 45%, with 30% reporting chronic cough, 21% reporting frequent phlegm production and wheezing and 26% reporting dyspnoea [14]. Similarly, in Cameroonian HIV patients, Pefura-Yone *et al* reported the prevalence of any respiratory symptom to be 47.5% while that of cough and dyspnoea were 25.5% and 36.4% respectively [16]. Contrary to the finding of our study, the prevalence of respiratory symptoms reported in Nigerian HIV patients was low (17%) [13] and HIV patients in South Africa had <4% of any respiratory symptoms [17]. Systematic review and metanalysis of studies conducted in developed countries have similarly reported a high prevalence of respiratory symptoms irrespective of access to HAART and high viremic control [7]. In our study, we found that in addition to having HIV, increasing age, exposure to medium-high levels of biomass fumes, unemployment and no/basic education were predictors of respiratory symptoms. Respiratory symptoms are known to be common with increasing age due to prolonged exposure to environmental toxins and decreased physiological capacity [10]. Our population has a low smoking burden but high biomass exposure and environmental pollution common in the peri-urban area might be responsible for the high prevalence of respiratory symptoms [29]. With the popularity of spirometry for screening airway obstruction, respiratory symptoms are being given less attention. However, the symptoms experienced by HIV patients indicates functional status and quality of life, even if they correlate poorly with spirometry [7].

### Limitations of the study

This study was conducted in a single primary healthcare facility in peri-urban Ghana, and hence, the findings may differ from the urban and rural populations, as well as other healthcare facilities in a different stratum. We cannot infer causation from the findings of the study due to the cross-sectional design. It may be possible that some of our participants may have had abnormal spirometry, especially RSP, before HIV infection. The data on chronic respiratory symptoms may be affected by recall bias, which may lead to the misclassification of some study participants. In this study, airway obstruction was defined by single spirometry. It has been that about 20% and 28% of patients initially diagnosed with OSP reversed to normal pattern after one and two years, respectively. Among patients diagnosed with normal spirometric patterns, 11% and 17% recorded OSP after one and two years, respectively [31]. Therefore, with single spirometry as in this study, we cautiously avoided categorising our study participants as having COPD to avoid this miscalssification. In addition, we used the GLI reference equation based on African American population for computing prediction values and LLN for spirometric values. The GLI equation is reported to significantly underestimate FEV1 and FVC by 9% in the Cameroonian population [32], implying that the performance of GLI equations in the SSA adult population may be limited. Therefore, there is a need for deriving locally reference values from the Ghanaian population for accurate interpretation of spirometric values.

### Conclusion

In conclusion, our study has shown that PLWH without any recent pulmonary infection in a peri-urban area of Ghana, the prevalence of OSP was similar among HAART-treated HIV and HAART-naïve HIV patients compared to the non-HIV control. However, the prevalence of RSP is higher in HIV patients on HAART treatment. This study has also shed light on the various predictors of spirometric abnormalities with the hope that future studies may investigate the possibility of using interventions targeted at the various risk factors to reduce pulmonary dysfunction in HIV patients.

## Data Availability

The dataset supporting the conclusion of this study is available for systematic review and meta-analysis upon request from the corresponding author.

## Abbreviations

SSA: sub-Saharan Africa
HIV: human immunodeficiency virus
PLWH: people living with HIV
HAART: highly active antiretroviral therapy
FEV1: forced expiratory volume in 1 s
FVC: forced vital capacity
LLN: lower limit of normal
OSP: obstructive spirometric pattern
RSP: restrictive spirometric patterns
GLI: Global Lung Initiative.

## Supporting information

Table S1. Determinants of obstructive lung pattern in HAART-treated HIV patients

Table S2. Determinants of obstructive lung pattern in HAART-naïve HIV patients

Table S3. Determinants of obstructive lung pattern in treatment non-HIV controls

Table S4. Determinants of restrictive lung pattern in HAART-treated HIV patients

Table S5. Determinants of restrictive lung pattern in HAART-naïve HIV patients

Table S6. Determinants of restrictive lung pattern in non-HIV controls

Table S7. Respiratory symptoms and spirometric abnormalities among HAART-treated HIV patients

## Acknowledgements

Our sincere thanks go to the staff and patients of Atua Government Hospital who assisted in data collection, especially Mr Samuel Essel and Mrs Nneka Essel.

## Funding

There was no funding for this study.

### Availability of data and materials

The dataset supporting the conclusion of this study is available for systematic review and meta-analysis upon request.

## Contributions

KY contributed to the conception, design, data analysis, and drafting of the manuscript and bears the primary responsibility for the content of the manuscript.

LM collected the data and revised the manuscript.

KBA was involved in the supervision and revision of the manuscript.

All the authors read and approved the content of the manuscript.

## Ethics approval and consent to participate

The study was conducted in conformity with the Helsinki Declaration on Human Experimentation, 1964 with subsequent revisions, latest Seoul, October 2008. Ethical approval for the study was granted by the Ethics and Protocol Review Committee of the College of Health Science, University of Ghana (Protocol ID number: KBTH-IRB /00131/2019). All the participants provided voluntary written consent before being recruited into the study.

## Consent for publication

Not applicable

## Reference

1. James SL, Abate D, Abate KH, Abay SM, Abbafati C, Abbasi N, Abbastabar H, Abd-Allah F, Abdela J, Abdelalim A: Global, regional, and national incidence, prevalence, and years lived with disability for 354 diseases and injuries for 195 countries and territories, 1990–2017: a systematic analysis for the Global Burden of Disease Study 2017. The Lancet 2018, 392(10159):1789–1858.

2. Roth GA, Abate D, Abate KH, Abay SM, Abbafati C, Abbasi N, Abbastabar H, Abd-Allah F, Abdela J, Abdelalim A: Global, regional, and national age-sex-specific mortality for 282 causes of death in 195 countries and territories, 1980–2017: a systematic analysis for the Global Burden of Disease Study 2017. The Lancet 2018, 392(10159):1736–1788.

3. Ghana AIDS Commission: National and Sub-National HIV and AIDS Estimates and Projections 2020 Report. In., vol. 2020. Accra: Ghana AIDS Commision; 2021.

4. Teeraananchai S, Kerr S, Amin J, Ruxrungtham K, Law M: Life expectancy of HIV-positive people after starting combination antiretroviral therapy: a meta-analysis. HIV medicine 2017, 18(4):256–266.

5. Yang H-Y, Beymer MR, Suen S-c: Chronic Disease Onset Among People Living with HIV and AIDS in a Large Private Insurance Claims Dataset. Scientific Reports 2019, 9(1):18514.

6. Bigna JJ, Kenne AM, Asangbeh SL, Sibetcheu AT: Prevalence of chronic obstructive pulmonary disease in the global population with HIV: a systematic review and meta-analysis. The Lancet Global Health 2018, 6(2):e193–e202.

7. Brown J, Roy A, Harris R, Filson S, Johnson M, Abubakar I, Lipman M: Respiratory symptoms in people living with HIV and the effect of antiretroviral therapy: a systematic review and meta-analysis. Thorax 2017, 72(4):355–366.

8. Drummond MB, Huang L, Diaz PT, Kirk GD, Kleerup EC, Morris A, Rom W, Weiden MD, Zhao E, Thompson B et al: Factors associated with abnormal spirometry among HIV-infected individuals. Aids 2015, 29(13):1691–1700.

9. Gupte AN, Wong ML, Msandiwa R, Barnes GL, Golub J, Chaisson RE, Hoffmann CJ, Martinson NA: Factors associated with pulmonary impairment in HIV-infected South African adults. PLOS ONE 2017, 12(9):e0184530.

10. Vaz Fragoso CA, Gill TM: Respiratory Impairment and the Aging Lung: A Novel Paradigm for Assessing Pulmonary Function. The Journals of Gerontology: Series A 2011, 67A(3):264–275.

11. Venkateshiah SB, Ioachimescu OC, McCarthy K, Stoller JK: The Utility of Spirometry in Diagnosing Pulmonary Restriction. Lung 2008, 186(1):19–25.

12. Guerra S, Sherrill DL, Venker C, Halonen M, Martinez FD: Chronic Restrictive Lung Pattern Is Associated with Increased Mortality Risk. In: American Thoracic Society 2009 International Conference. vol. C34. COPD PHENOTYPES. San Diego Convention centre: American Journal of Respiratory and Critical Care Medicine; 2009: A4325.

13. Akanbi MO, Taiwo BO, Achenbach CJ, Ozoh OB, Obaseki DO, Sule H, Agbaji OO, Ukoli CO: HIV Associated Chronic Obstructive Pulmonary Disease in Nigeria. J AIDS Clin Res 2015, 6(5).

14. Ddungu A, Semitala FC, Castelnuovo B, Sekaggya-Wiltshire C, Worodria W, Kirenga BJ: Chronic obstructive pulmonary disease prevalence and associated factors in an urban HIV clinic in a low income country. PLOS ONE 2021, 16(8):e0256121.

15. Kayongo A, Wosu AC, Naz T, Nassali F, Kalyesubula R, Kirenga B, Wise RA, Siddharthan T, Checkley W: Chronic Obstructive Pulmonary Disease Prevalence and Associated Factors in a Setting of Well-Controlled HIV, A Cross-Sectional Study. COPD: Journal of Chronic Obstructive Pulmonary Disease 2020, 17(3):297–305.

16. Pefura-Yone EW, Fodjeu G, kengne AP, Roche N, Kuaban C: Prevalence and determinants of chronic obstructive pulmonary disease in HIV infected patients in an African country with low level of tobacco smoking. Respiratory Medicine 2015, 109(2):247–254.

17. van Riel SE, Klipstein-Grobusch K, Barth RE, Grobbee DE, Feldman C, Shaddock E, Stacey SL, Venter WDF, Vos AG: Predictors of impaired pulmonary function in people living with HIV in an urban African setting. South Afr J HIV Med 2021, 22(1):1252.

18. Ferris BG: Epidemiology Standardization Project (American Thoracic Society). Am Rev Respir Dis 1978, 118(6 Pt 2):1–120.

19. Graham BL, Steenbruggen I, Miller MR, Barjaktarevic IZ, Cooper BG, Hall GL, Hallstrand TS, Kaminsky DA, McCarthy K, McCormack MC et al: Standardization of Spirometry 2019 Update. An Official American Thoracic Society and European Respiratory Society Technical Statement. American Journal of Respiratory and Critical Care Medicine 2019, 200(8):e70–e88.

20. Quanjer PH, Stanojevic S, Cole TJ, Baur X, Hall GL, Culver BH, Enright PL, Hankinson JL, Ip MSM, Zheng J et al: Multi-ethnic reference values for spirometry for the 3–95-yr age range: the global lung function 2012 equations. European Respiratory Journal 2012, 40(6):1324–1343.

21. van Gemert F, Kirenga B, Chavannes N, Kamya M, Luzige S, Musinguzi P, Turyagaruka J, Jones R, Tsiligianni I, Williams S et al: Prevalence of chronic obstructive pulmonary disease and associated risk factors in Uganda (FRESH AIR Uganda): a prospective cross-sectional observational study. Lancet Glob Health 2015, 3(1):e44–51.

22. Schultz K, D’Aquino LC, Soares MR, Gimenez A, Pereira CAdC: Lung volumes and airway resistance in patients with a possible restrictive pattern on spirometry. Jornal Brasileiro de Pneumologia 2016, 42:341–347.

23. Vaz Fragoso CA, Van Ness PH, Murphy TE, McAvay GJ: Spirometric impairments, cardiovascular outcomes, and noncardiovascular death in older persons. Respiratory Medicine 2018, 137:40–47.

24. Siddharthan T, Grigsby M, Miele CH, Bernabe-Ortiz A, Miranda JJ, Gilman RH, Wise RA, Porter JC, Hurst JR, Checkley W: Prevalence and risk factors of restrictive spirometry in a cohort of Peruvian adults. Int J Tuberc Lung Dis 2017, 21(9):1062–1068.

25. Hyatt RE, Cowl CT, Bjoraker JA, Scanlon PD: Conditions Associated With an Abnormal Nonspecific Pattern of Pulmonary Function Tests. CHEST 2009, 135(2):419–424.

26. Aryal S, Diaz-Guzman E, Mannino DM: COPD and gender differences: an update. Translational Research 2013, 162(4):208–218.

27. Camp PG, Goring SM: Gender and the Diagnosis, Management, and Surveillance of Chronic Obstructive Pulmonary Disease. Proceedings of the American Thoracic Society 2007, 4(8):686–691.

28. Nketiah-Amponsah E, Afful-Mensah G, Ampaw S: Determinants of cigarette smoking and smoking intensity among adult males in Ghana. BMC Public Health 2018, 18(1):941.

29. Torres-Duque C, Maldonado D, Pérez-Padilla R, Ezzati M, Viegi G: Biomass Fuels and Respiratory Diseases. Proceedings of the American Thoracic Society 2008, 5(5):577–590.

30. Silva R, Oyarzún M, Olloquequi J: Pathogenic Mechanisms in Chronic Obstructive Pulmonary Disease Due to Biomass Smoke Exposure. Archivos de Bronconeumología (English Edition) 2015, 51(6):285–292.

31. Schermer TR, Robberts B, Crockett AJ, Thoonen BP, Lucas A, Grootens J, Smeele IJ, Thamrin C, Reddel HK: Should the diagnosis of COPD be based on a single spirometry test? npj Primary Care Respiratory Medicine 2016, 26(1):16059.

32. Pefura-Yone EW, Balkissou AD, Poka-Mayap V, Djenabou A, Massongo M, Ofimboudem NA, Mayoh-Nguemfo CF, Tsala AG, Hadjara H, Amougou F: Spirometric reference equations for Cameroonians aged 4 to 89 years derived using lambda, mu, sigma (LMS) method. BMC Pulm Med 2021, 21(1):344.

